# Delayed clearance of SARS-CoV2 in male compared to female patients: High ACE2 expression in testes suggests possible existence of gender-specific viral reservoirs

**DOI:** 10.1101/2020.04.16.20060566

**Authors:** Aditi Shastri, Justin Wheat, Sachee Agrawal, Nirjhar Chaterjee, Kith Pradhan, Mendel Goldfinger, Noah Kornblum, Ulrich Steidl, Amit Verma, Jayanthi Shastri

## Abstract

The novel coronavirus SARS-CoV2 has been observed to cause a higher incidence and greater severity of disease in males, as seen in multiple cohorts across the globe. The reasons for gender disparity in disease severity is unclear and can be due to host factors. To determine whether males have delayed viral clearance after infection, we evaluated the time to clearance in symptomatic patients tested by serial oropharyngeal/nasopharyngeal swabs followed by RT-PCR at a reference lab in Mumbai, India. A total of 68 subjects with median age of 37 years (3-75 range) were examined and included 48 (71%) males and 20 (29%) females. We observed that females were able to achieve viral clearance significantly earlier than males, with a median difference of 2 days in achieving a negative PCR result (P value = 0.038). Furthermore, examination of 3 families with both male and female patients followed serially, demonstrated that female members of the same household cleared the SARS-CoV2 infection earlier in each family. To determine reasons for delayed clearance in males, we examined the expression patterns of the SARS-CoV2 receptor, Angiotensin-converting enzyme 2 (ACE2), in tissue specific repositories. We observed that the testes was one of the highest sites of ACE2 expression in 3 independent RNA expression databases (Human Protein Atlas, FAMTOM5 and GETx). ACE2 was also determined to be highly expressed in testicular cells at the protein levels. Interestingly, very little expression of ACE2 was seen in ovarian tissue. Taken together, these observations demonstrate for the first time that male subjects have delayed viral clearance of SARS-CoV2. High expression of ACE2 in testes raises the possibility that testicular viral reservoirs may play a role in viral persistence in males and should be further investigated.

## Introduction

The novel coronavirus SARS-CoV2, the causative infectious agent of the COVID19 pandemic, represents a significant risk to global health, health care infrastructure, the global financial markets, and social stability. As coronaviruses rarely transfer from their zoonotic species to humans and cause serious disease, there remain many questions related to how these viruses interact with their human host to propagate infection which is causative of the resultant morbidity and mortality.

Multiple early studies have demonstrated a gender imbalance in disease severity and duration, with both the incidence of disease and morbidity in men being more than double that of women.^1,2^ This is similar to prior data regarding disease severity with the SARS-CoV1 outbreak in the early 2000s and the MERS epidemic.^3,4^ While some of these effects have been attributed to the known higher rates of cardiovascular comorbidities and associated risk factors such as hypertension, smoking, and coronary artery disease that are more predominant in men than women, a molecular mechanism for this increased propensity towards causing gender-specific morbidity is still speculative. ^5-7^

## Methods

The clinical protocol was IRB approved by the Kasturba Hospital for Infectious Diseases Institutional Review Board on April 7, 2020. The first positive case for SARS-CoV 2 was detected in Mumbai on March 11th 2020. Patients with suspected symptoms were admitted to the isolation ward at the Kasturba Hospital & monitored. Nasopharyngeal/ orophryngeal swabs collected at the time of diagnosis were analyzed by RT-PCR for positivity/negativity per the Berlin protocol.^8^ Subsequent swabs were collected at approximately 48 hours intervals until the swab test was documented to be negative. Persons residing in the same household as the index case as well as contacts were also tested. A log rank test was used to compare the difference in viral clearance between males and females and is represented in Figure 1.B using Kaplan-Meier curves. Mean & median times towards viral clearance were also calculated for each gender. Statistical calculations and analysis were performed in the R version 3.6.1 environment.

**Fig 1:**
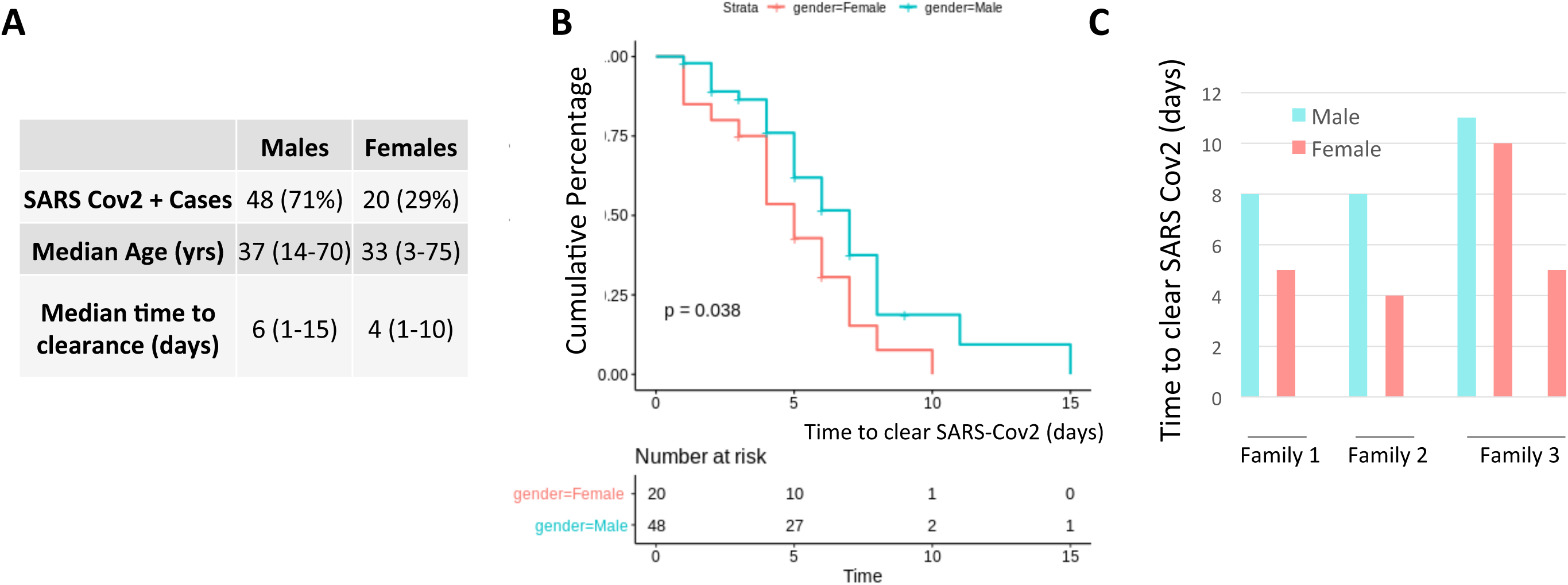
Female subjects clear SARS Cov2 faster than males: A: Table showing demographics and patient characteristics B: Kaplan Meir curves showing rates of viral clearance in male and female subjects. Log rank p value= 0.038 C: Time to viral clearance shown for 3 families with SARS-CoV2 positivity

Three RNA and protein expression datasets (Human Protein Atlas, FAMTOM5 and GETx) were subsequently queried to quantify the expression of the Angiotensin-converting enzyme 2 (ACE2) which is the receptor for SAR-CoV2 entry into human tissue. mRNA as well as protein expression was queried in each organ.

## Results and Discussion

### Delayed clearance of SARS-CoV2 is seen in males

To test whether males have delayed clearance, we examined patterns in patients that were symptomatic and tested positive for SARS-CoV2 by swabs followed by RT-PCR at the Molecular Diagnostics Laboratory at the Kasturba Hospital for Infectious Diseases in Mumbai, India. A total of 68 subjects with median age of 37 years (3-75 range) were examined. The subjects included 48 (71%) males and 20 (29%) females (Fig 1A). SARS-CoV2 clearance was determined by the number of days it took to achieve a persistently negative test by RT-PCR. The procedure involved was that serial oropharyngeal/nasopharyngeal swabbing till negative results were obtained. We observed that females were able to achieve viral clearance significantly earlier than males (Fig 1B, Log Rank P value = 0.038). Female subjects achieved viral clearance at a median of 4 days (range 1-10 days) vs a median of 6 days (range 1-15 days) in male subjects (Fig 1A,B).

As the population was composed of heterogeneous subjects, we next identified 3 families (with all family members residing in the same household) with a recent positive swab for SARS-CoV2. In all 3 families, the female members cleared the SARS-COV2 infection earlier than the male members of the same family (Fig 1C). These data show for the first time that males clear the virus later than females.

### Testes express high levels of ACE2 receptor

SARS-CoV2 enters cells via a complex, multiple-step process which begins with interaction of the viral surface spike protein with ACE2.^9,10^ ACE2 is a plasma membrane bound monocarboxypeptidase that converts angiotensin II to angiotensin 1-7 (Ang 1-7), which antagonizes the hypertensive effects of Angiotensin I via stimulation of the MAS receptor.^11^ We evaluated ACE2 expression at the mRNA and protein levels using tissue specific databases. We observed that ACE2 is expressed at high levels in the lungs, GI tract, myocardium, testes, and brush border cells of the renal proximal tubule at the mRNA levels (Fig 2A). Interestingly, the testes was one of the sites with highest ACE2 expression in 3 independent RNA expression databases (Human Protein Atlas, FAMTOM5 and GETx) (Fig 2B), consistent with prior reports.^12-14^ ACE2 was also determined to be highly expressed in testicular cells at the protein levels (Fig 2D, E). Interestingly, very little expression was seen in ovarian tissue (Fig 2C,F). These data raise the possibility that SARS-CoV2 may bind to testicular ACE2 and the testes may serve as a male specific viral reservoir. Consistent with this hypothesis, a very recent study demonstrated testicular gonadal loss-of-function in SARS-CoV2 patients, suggesting possible damage to testicular cells during infection.^15^

**Fig 2:**
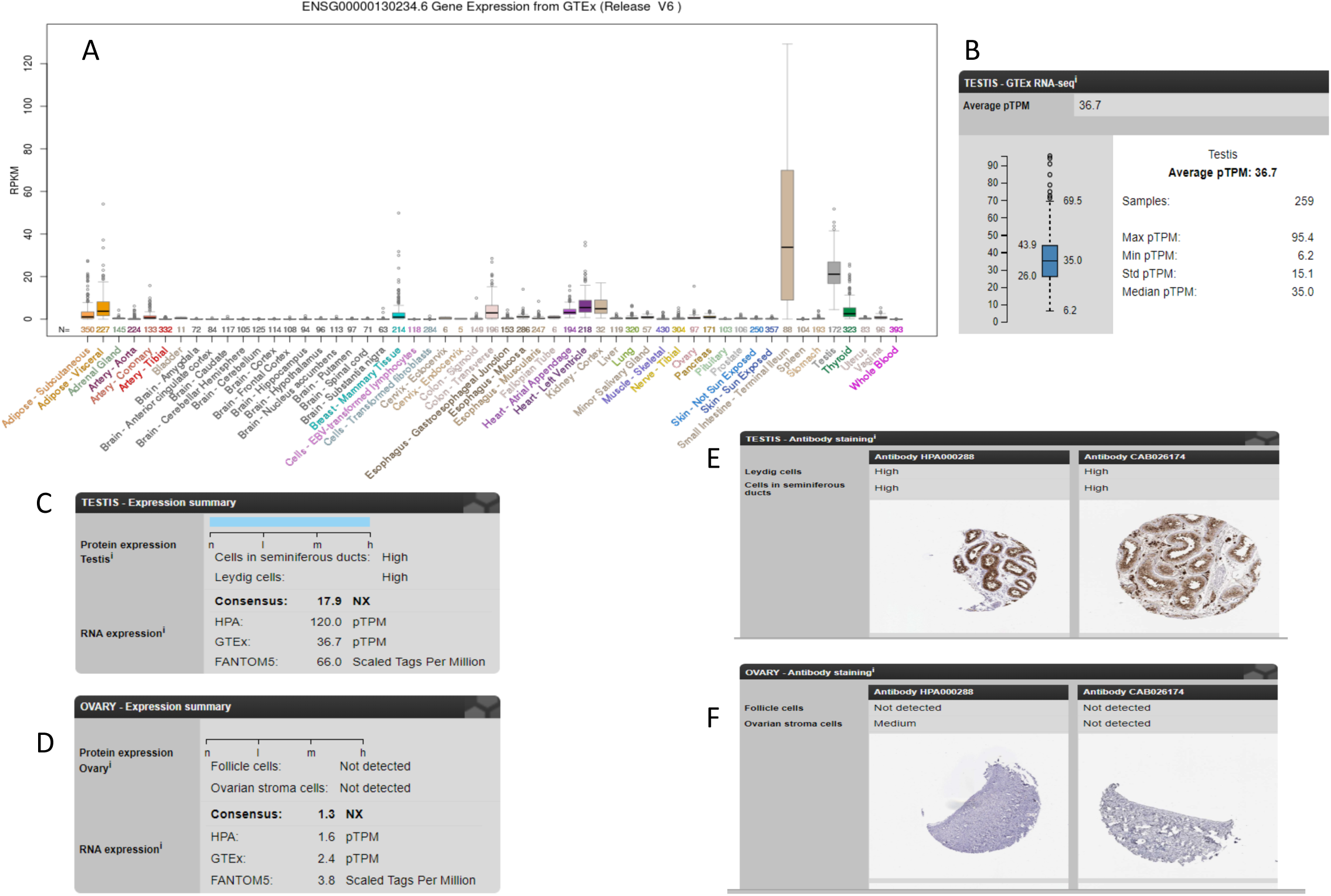
ACE2 is highly expressed in testes: A, B: mRNA expression data from GETX RNA-seq repository shows that testicular tissue has a high expression of ACE2 C, D: ACE2 expression at the protein level and mRNA level is seen in 3 independent databases (Human Protein Atlas (HPA), FANTOM5 and GETx) in testes, and is not detected in ovarian tissues E,F: Representative tissue sections showing high ACE2 protein expression using two different antibodies in testes (Human Protein Atlas). ACE2 is not detected in ovarian tissues.

Taken together, these observations demonstrate that male subjects have delayed viral clearance. High expression of ACE2 RNA and protein in testes leads to the hypothesis that testicular viral reservoirs may exist and play a role in viral persistence, and should be further investigated by larger clinical studies. This hypothesis may have important implications for understanding the transmission and persistence of SARS-CoV2 virus in humans, and in a gender-specific manner.

## Data Availability

Data highlighted in the manuscript is available publicly in Human Protein Atlas, FAMTOM5 and GETx. The patient data reported is confidential and protected by an IRB approved protocol.

